# STRENGTH OF ASSOCIATION BETWEEN GENERALIZED/NONSPECIFIC COVID-19 SIGNS & SYMPTOMS WITH SARS-COV 2 SPECIFIC ORF, N, E GENES IDENTIFIED THROUGH REAL TIME PCR

**DOI:** 10.1101/2022.11.16.22282408

**Authors:** Muhammad Arif, Muhammad Abdullah, Sheikh Ahmed, Ehsan Ahmed Larik, Ujala Naseer Baloch, Zakir Hussain, Mirza Zeeshan Iqbal Baig

## Abstract

**BACKGROUND:** Constant mutation in the SARS-COV2 virus genetic material is resulting in the appearance of new variants frequently hence the overall virulence, treatment resistance, replication modalities, transmissions rates and COVID-19 signs & symptoms are all changing regularly.

**METHODOLOGY:** From 1 January 2021 to 30 August 2022, the clinical lab at Fatima Jinnah General & Chest Hospital Quetta, Balochistan, determined a total of 3375 individuals to be COVID-19 positive because RT-PCR detected ORF, N, and E genes or their various Bi & Tri combinations in their samples. A questionnaire-based interview was conducted with each participant during sample collection. Body temperature more than 37^0^c was recorded as Fever/Chill. Age, Comorbidities, A-symptomatic individuals & Vaccination status were all neglected during this study. Frequency tables were generated using MS-excel 2016, while Odds ratios were calculated using Chi-square test of association whereby 2×2 contingency tables between Mono, Bi & Tri combinations for ORF, N & E genes were cross associated with various generalized nonspecific COVID -19 signs and symptoms using Epi-info software. Absence of Genetic sequencing was the major limitation.

**RESULTS:** The study showed that individually the presence of **ORF gene** was found to be strongly associated **“ Shortness of Breath/Difficulty in Breathing”, Diarrhea, Head ache & Vomitting**. While the presence of **N-gene** was found to be strongly associated with **Loss of smell & taste, Head ache**,**Presistant Chest Pain & Bluish lips/Face**. Where as the presence of **E-gene** was found to be strongly associated with **Cough, Shortness of breath/ Difficulty in breathing, Sore throat, Diarrhea, Head ache & Laziness**. In addition, the study also found that different Bi & Tri combinations of ORF, N & E genes in a COVID-19 positive patient expressed generalized non-specific COVID-19 signs & symptoms differently.

**DISCUSSION & CONCLUSION:** The presence of various SARS-COV2 genetic markers significantly alters the clinical presentation of COVID-19.

## INTRODUCTION

There were 41,409 people in all documented, from at least 23 different nations, with 26 different clinical presentations. Six symptoms—fever (58.66%), cough (54.52%), dyspnea (30.82%), malaise (29.75%), weariness (28.16%), and sputum/secretion (25.33%)—had a general prevalence more than or equal to 25%. Other prevalent symptoms included headache (12.17%), chest discomfort (11.49%), diarrhea (9.59%), sneezing (14.71%), sore throat (14.41%), rhinitis (14.29%), goosebumps (13.49%), dermatological signs (20.45%), anorexia (20.26%), myalgia (16.9%), and rhinitis. The manifestations of dermatology were only documented in one study. Hemoptysis was the least common indication or symptom (1.65%). The three most common symptoms in trials involving more than 100 patients were dyspnea (30.64%), cough (54.21%), and fever (57.93%) ^[1-13]^.

Certain symptoms, such as dyspnea, fever, cough, and headache, are generally nonspecific for SARS-CoV2. Patients who are asymptomatic may have mild forms of infection, while others may have severe pneumonia that can be fatal ^[2-3]^.

The initial symptoms of the illness were the trio of fever, coughing, and shortness of breath. Later, the US Center for Disease Control and Prevention (CDC) expanded this list to include chills, headache, sore throat, muscle discomfort, and loss of taste or smell (neurological manifestations) ^[4-15]^.

Any possible association between generalized nonspecific COVID-19 signs and symptoms with SARS-COV 2 specific ORF, N & E genes would open up a new door to research in future.

### LITERATURE REVIEW

It is certain that age manifest the clinical sings & symptoms of COVID-19 differently. According to one study, people over 60 have more bilateral lobe lesions, greater levels of inflammatory markers, and higher blood urea nitrogen levels. Patients who are older than 60 have a higher risk of respiratory failure and prolonged illness courses. The intensity is, however, less severe in people under 60 ^[5-14]^.

According to one additional study that claims a total of 72,314 verified cases in China the majority of the patients (87%) are between the ages of 30 and 79. There were no fatalities among those under the age of nine. However, the case-fatality rate (CFR) is 8.0% for those aged 70 to 79 and 14.8% for people 80 years of age and older. The CFR is 10.5, 7.3, 6.3, 6.0, and 5.6%, respectively, for patients with various concomitant diseases, including cancer, chronic respiratory disease, diabetes, cardiovascular disease, and hypertension. According to these findings, COVID-19 patients with comorbid conditions have higher fatality rates than those who don’t have underlying diseases. ^[6-15]^

Coronaviruses have genomes that range in size from 26 to 32 kilobases and have a variety of open reading frames (ORFs) ^[7]^. Coronaviruses have a varying number of open reading frames in their genomes, which range in size from 26 to 32 kilobases (ORFs). The spike surface glycoprotein (S), small envelope protein (E), matrix protein (M), and nucleocapsid protein (N) are the four major structural proteins and the eight accessory proteins (3a, 3b, p6, 7a, 7b, 8b, 9b, and orf14) are situated in the 3′-terminus of the SARS-CoV-2 genome ^[8-16-17]^.

The SARS-CoV-2 was found to be more related to two SARS-like bat CoVs from Zhoushan in eastern China, bat-SL-CoVZC45 and bat-SL-CoVZXC21, than to the SARS-CoV and the MERS-CoV, according to analysis of the genome from the samples of nine patients. Laboratory specific detection Next-generation sequencing or real-time reverse transcriptase-polymerase chain reaction (RT-PCR) techniques for the SARS-CoV-2 virus were developed as a result of the isolation of the causative agent and determination of its partial genomic sequence ^[9-18-19-20]^.

The open reading frame (ORF) segments in the SARS COV 2 virus genome encode structural and non-structural proteins ^[8]^. The ORF1a/b gene in SARS COV 2 nucleic acid is used for diagnostics by RT PCR. It produces non-structural proteins (nsp1–16), which are necessary for the viral genome’s maintenance and replication apparatus. Adaptive mutations in ORF1a/b may boost viral replication or increase treatment resistance, hence increasing virulence ^[10}^. The “N” genome encodes the “N” structural protein which participates in a number of viral genome-related functions, such as viral genome signaling, viral replication, and host cell immunity to viral infections ^[11]^. Similarly, the “E” genome encodes for the “E” structural protein Through interactions with the host cell membrane protein, the E protein participates in the viral growth and maturation stages ^[12-21-22-23-24-25]^.

From the very first case the SARS-COV 2 infection, a continuous mutation is reported in the virus as a result of which new variants popup frequently hence the overall virulence, treatment resistance, replication modalities & transmissions rates are all changing regularly hence in these circumstances it is of great importance to frequently monitor the symptomology of COVID-19. Currently no published literature has assessed the relationship of COVID-19 symptomology (i.e. *COUGH, SHORTNESSS OF BREATH/DIFFICULTY IN BREATHING, Fever/Chills, NEW MUSCLE/BODY ACHE, SORE THROAT, LOSS OF SMELL & TASTE, DIARRHEA, HEAD ACHE, NAUSEA, VOMITTING, RUNNY NOSE, PERSISTANT CHEST PAIN, LAZINESS, BLUISH LIPS/FACE*) with the *<u>ORF, N & E</u>* genes which are few of the diagnostic markers assessed for SARS COV-2 infection assessed during real-time PCR test.

## METHODOLOGY

From 1 January 2021 to 30 August 2022, the clinical lab at Fatima Jinnah General & Chest Hospital Quetta, Balochistan, determined a total of 3375 individuals to be COVID-19 positive because RT-PCR detected ORF, N, and E genes or their various Bi & Tri combinations in their samples. A questionnaire-based interview was conducted with each participant during sample collection. Body temperature more than 37^0^c was recorded as Fever/Chill. Age, Comorbidities, A-symptomatic individuals & Vaccination status were all neglected during this study. Frequency tables were generated using MS-excel 2016, while Odds ratios were calculated using Chi-square test of association whereby 2×2 contingency tables between Mono, Bi & Tri combinations for ORF, N & E genes were cross associated with various generalized nonspecific COVID -19 signs and symptoms using Epi-info software. Absence of Genetic sequencing was the major limitation.

## RESULTS

From 01 January 2021 till 30^th^ August 2022 a total of 3375 local residents across Balochistan were declared to be COVID-19 positive by the RT-PCR. 48% (n=1620) individuals out of 3375 study participants were found to be positive for all the three major diagnostic markers of SARS-COV 2 (i.e. ORF + N + E). Similarly, the RT-PCR reports of 12% (n=405) study participants were positive for only N & E (N + E) genes together. Moreover 13% (n= 438) study participants were positive only for ORF & N (ORF + N) genes together. While 10% (n=337) study participants were positive only for ORF & E (ORF+E) genes combination. It was also seen that 08% (n=270) study participants were positive only for ORF gene, 06% (n=203) study participants expressed only “N” gene while only 03% (n=102) of the study participants were positive only for “E” gene only as shown below:

**Fig No: 01.**
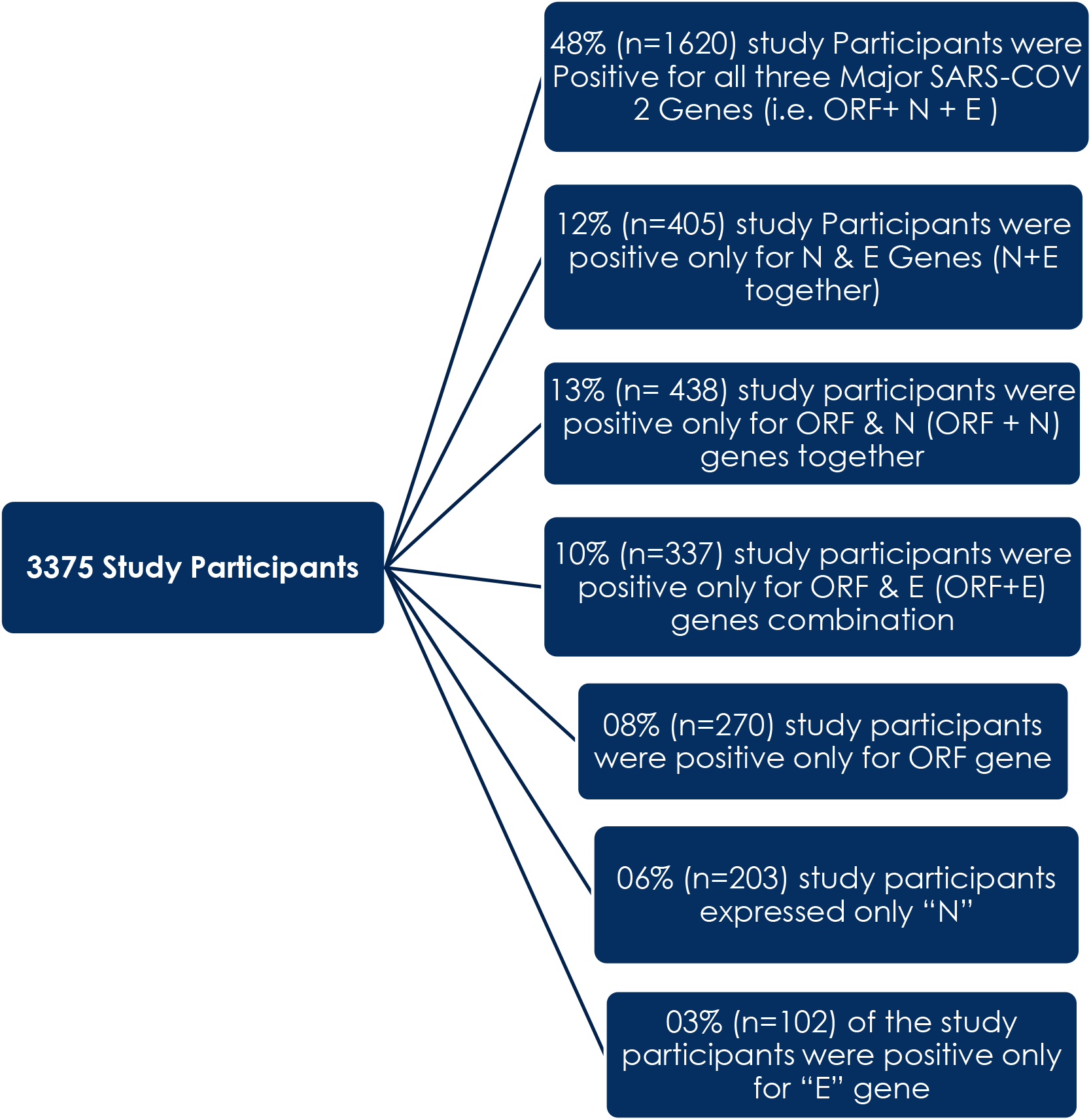
Sub classification of the study participants based on results of RT-PCR reports.

### VIRAL LOAD OF SARS-COV 2 AMONG STUDY PARTICIPANTS

If a sample gets positive and show presence of ORF, N & E genes in any combination on lesser RT-PCR cycles i.e. below 21 cycles is generally believed to possess high viral load while if a sample becomes positive at 21 or beyond RT-PCR cycles it is beloved to have low vial load. In this Study out of 3375 study participants 57%(n=1915) were positive with high viral load while rest of the 42% (n=1460) were found to be positive with low viral load as shown in the following chart:

**Chart no 01:**
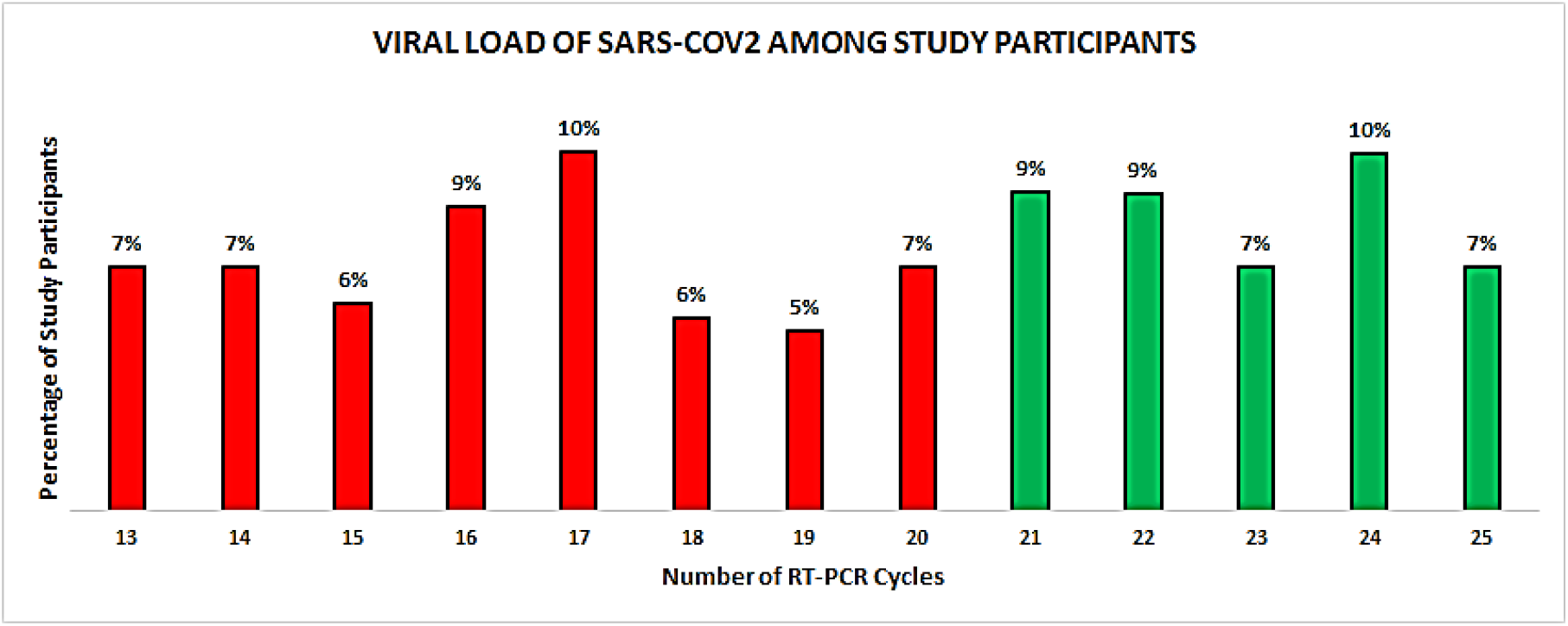
Viral load of study participants.

The following table further summarizes the above chart and is showing cycle wise positivity rate:

**Table No. 01:**
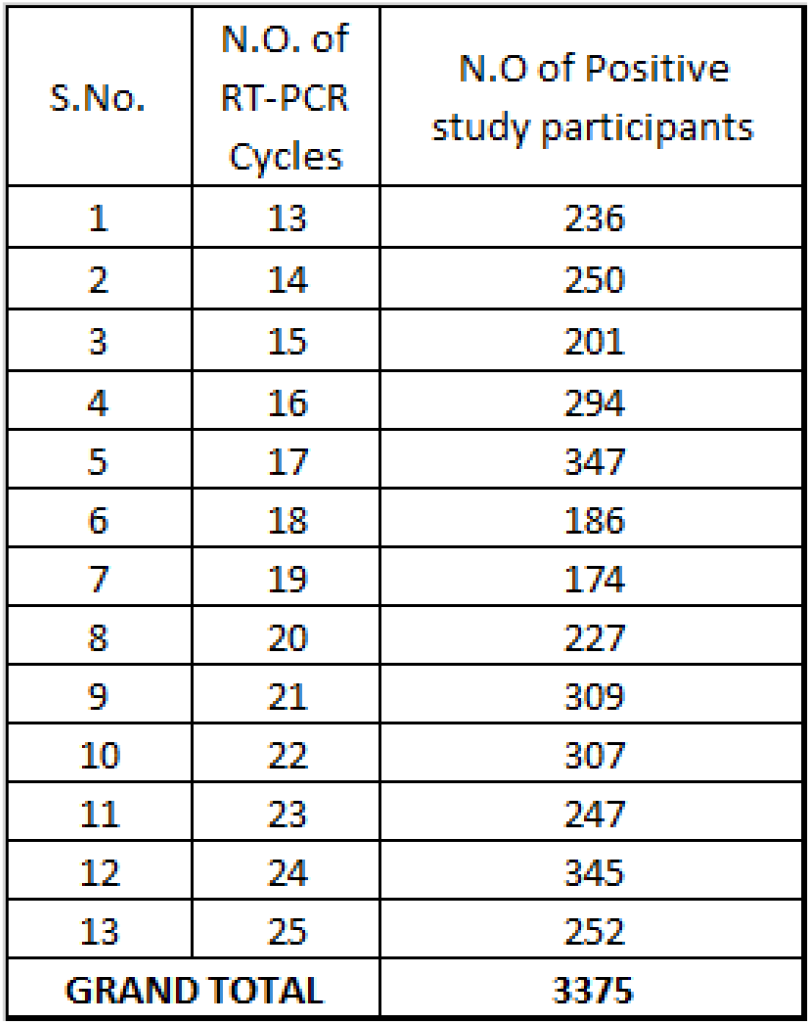
Positivity rate against RT-PCR Cycles.

### FREQUENCY & PERCENTAGES OF THE GENERALIZED/NON SPECIFIC SIGN AND SYMPTOMS AMONG THE STUDY PARTICIPANTS

The following table summarizes the overall findings:

**Table No.02:** Overall summary of the major findings.

FREQUENCY & PERCENTAGES OF THE GENERALIZED/NON SPECIFIC SIGN AND SYMPTOMS AMONG THE STUDY PARTICIPANTS:
| Sub classification of study participants on the bases of RT-PCR Report | SARS_COV2 (ORF) Gene | SARS_COV2 (N) Gene | SARS_COV2 (E) Gene | Total Number of Positive Samples | Cough |  | Shortness of Breath/ Difficulty in Breathing |  | Fever/Chill |  | New Mucle/Bod y aches |  | Sore Throat |  | Loss of Smell & taste |  | Diarrhoea |  | Head Ache |  | Nausea |  | Vomitting |  | Runny Nose |  | Persistant chest pain or pressure |  | Laziness |  | Blush lips or face |  |
| --- | --- | --- | --- | --- | --- | --- | --- | --- | --- | --- | --- | --- | --- | --- | --- | --- | --- | --- | --- | --- | --- | --- | --- | --- | --- | --- | --- | --- | --- | --- | --- | --- |
|  |  |  |  |  | Yes | No | Yes | No | Yes | No | Yes | No | Yes | No | Yes | No | Yes | No | Yes | No | Yes | No | Yes | No | Yes | No | Yes | No | Yes | No | Yes | No |
| All 03 Major Genes | + | + | + | 1620 (48%) | 513 | 1107 | 604 | 1016 | 1203 | 417 | 1310 | 310 | 519 | 1101 | 357 | 1263 | 284 | 1336 | 41 | 1579 | 842 | 778 | 1002 | 618 | 1338 | 282 | 1139 | 481 | 228 | 1392 | 695 | 925 |
| Different Combination of 02 Major Genes | - | + | + | 405 (12%) | 37 | 368 | 49 | 356 | 258 | 147 | 175 | 230 | 154 | 251 | 351 | 54 | 136 | 269 | 283 | 122 | 97 | 308 | 310 | 95 | 91 | 314 | 375 | 30 | 145 | 260 | 209 | 196 |
|  | + | + | - | 438 (13%) | 158 | 280 | 109 | 329 | 223 | 215 | 146 | 292 | 16 | 422 | 269 | 169 | 377 | 61 | 80 | 358 | 323 | 115 | 102 | 336 | 199 | 239 | 118 | 320 | 340 | 98 | 294 | 144 |
|  | + | - | + | 337 (10%) | 243 | 94 | 259 | 78 | 161 | 176 | 307 | 30 | 219 | 118 | 283 | 54 | 120 | 217 | 190 | 147 | 237 | 100 | 13 | 324 | 198 | 139 | 165 | 172 | 39 | 298 | 261 | 76 |
| Only single Major gene | + | - | - | 270 (08%) | 50 | 220 | 217 | 53 | 148 | 122 | 144 | 126 | 106 | 164 | 9 | 261 | 203 | 67 | 160 | 110 | 35 | 235 | 209 | 61 | 102 | 168 | 136 | 134 | 96 | 174 | 21 | 249 |
|  | - | + | - | 203 (06%) | 57 | 146 | 71 | 132 | 50 | 153 | 160 | 43 | 52 | 151 | 109 | 94 | 18 | 185 | 114 | 89 | 73 | 130 | 135 | 68 | 132 | 71 | 138 | 43 | 154 | 49 | 120 | 83 |
|  | - | - | + | 102 (03%) | 80 | 22 | 95 | 7 | 73 | 29 | 40 | 62 | 94 | 8 | 11 | 91 | 95 | 7 | 70 | 32 | 35 | 67 | 49 | 53 | 28 | 74 | 24 | 100 | 56 | 46 | 36 | 66 |
| GRAND TOTAL→ |  |  |  | 3375 | 1138 | 2237 | 1404 | 1971 | 2116 | 1259 | 2282 | 1093 | 1160 | 2215 | 1389 | 1986 | 1233 | 2142 | 938 | 2437 | 1642 | 1733 | 1820 | 1555 | 2088 | 1287 | 2095 | 1280 | 1058 | 2317 | 1636 | 1739 |
| PERCENTAGES → |  |  |  |  | 34% | 66% | 42% | 58% | 63% | 37% | 68% | 32% | 34% | 66% | 41% | 59% | 37% | 63% | 28% | 72% | 49% | 51% | 54% | 46% | 62% | 38% | 62% | 38% | 31% | 69% | 48% | 52% |

**Table No. 03:** Table adopted from the above table.

| S.NO. | Generalized/Non Specific COVID-19 Signs & Symptoms | Number of study Participant giving YES Responces | Percentages of study Participant giving YES Responces | Number of study Participant giving NO Responces | Percentage of study Participant giving NO Responces |
| --- | --- | --- | --- | --- | --- |
| 1 | COUGH | 1138 | 34% | 2237 | 66% |
| 2 | SHORTNESS OF BREATH/ DIFFICULTY IN BREATHING | 1404 | 42% | 1971 | 58% |
| 3 | Fever/Chills | 2116 | 63% | 1259 | 37% |
| 4 | NEW MUSCLE/BODY ACHE | 2282 | 68% | 1093 | 32% |
| 5 | SORE THROAT | 1160 | 34% | 2215 | 66% |
| 6 | LOSS OF SMELL & TASTE | 1389 | 41% | 1986 | 59% |
| 7 | DIARRHEA | 1233 | 37% | 2142 | 63% |
| 8 | HEAD ACHE | 938 | 28% | 2437 | 72% |
| 9 | NAUSEA | 1642 | 49% | 1733 | 51% |
| 10 | VOMITTING | 1820 | 54% | 1555 | 46% |
| 11 | RUNNY NOSE | 2088 | 62% | 1287 | 38% |
| 12 | PERSISTANT CHEST PAIN | 2095 | 62% | 1280 | 38% |
| 13 | LAZINESS | 1058 | 31% | 2317 | 69% |
| 14 | BLUISH LIPS/FACE | 1636 | 48% | 1739 | 52% |

34% of the participants has cough while 66% had no cough, similarly 42% of the study participants reported to have S.O.B while 58% did not have any S.O.B. Moreover 63% had fever/chill while 37% did not had. New muscular or body ach was reported by 68% individuals while 32% had no new muscular/body ache. Similarly, sore throat was reported by 34% of the study participants while 66% of the study participants did not report any sore throat symptom.

Loss of smell and taste was reported by 41% of the study participants while 59% did not report this symptom. Diarrhea was reported by 37% of the study participants while 63% did not report any diarrhea. 28% of the study participants reported headache while 72% did not report any such symptom. Nausea was reported by 49% of the study participants while 51% did not report this symptom. Similarly, 54% of the study participants reported vomiting while 46% did not report this symptom. Runny nose was reported by 62% of the study participants while 38% did not report any such symptom. 68% of the study participants reported persistent chest pain while 38% did not report any chest pain. Laziness was reported by 31% of the study participants and lastly Bluish lips/Face was reported by 48% of the study participants while 52% did not report any symptom.

### STRENGTH OF ASSOCIATION BETWEEN GENRALIZED/NON-SPECIFIC COVID-19 SINGS AND SYMPTOMS REPORTED BY ALL STUDY PARTICIPANTS WHOSE RT-PCR TEST WAS POSITIVE FOR ALL THREE SARS-COV 2 GENES (*i*.*e. ORF + N + E*) COMPARE TO OTHERS

The following two tables summarizes the overall findings as shown below:

**Table No.4:** Contingency 2×2 table between generalized/ non-specific COVID-19 Signs and symptoms reported by all the study participants whose RT-PCR report was positive for all the three SARS-COV 2 Genes (i.e. ORF + N + E) Compare to others.

|  |  | COUGH |  | SHORTNESS OF BREATH/DIFFICULTY IN BREATHING |  | Fever/Chills |  | NEW MUSCLE/ BODY ACHE |  | SORE THROAT |  | LOSS OF SMALL & TASTE |  | DIARRHEA |  |
| --- | --- | --- | --- | --- | --- | --- | --- | --- | --- | --- | --- | --- | --- | --- | --- |
|  |  | yes | No | yes | No | yes | No | yes | No | yes | No | yes | No | yes | No |
| SARS-COV-2 (ORF+N+E) GENES | + | 513 | 1107 | 604 | 1016 | 1203 | 417 | 1310 | 310 | 519 | 1101 | 357 | 1263 | 284 | 1336 |
|  | - | 625 | 1130 | 800 | 955 | 913 | 842 | 972 | 783 | 695 | 1060 | 1032 | 723 | 949 | 806 |

|  |  | HEAD ACHE |  | NAUSEA |  | VOMITTING |  | RUNNY NOSE |  | PERSISTANT CHEST PAIN |  | LAZINESS |  | BLUIISH LIPS/ FACE |  |
| --- | --- | --- | --- | --- | --- | --- | --- | --- | --- | --- | --- | --- | --- | --- | --- |
|  |  | yes | No | yes | No | yes | No | yes | No | yes | No | yes | No | yes | No |
| SARS-COV-2 (ORF+N+E) GENES | + | 41 | 1579 | 842 | 778 | 1002 | 618 | 1338 | 282 | 1139 | 481 | 228 | 1392 | 695 | 925 |
|  | - | 897 | 858 | 800 | 955 | 818 | 937 | 750 | 1005 | 956 | 799 | 830 | 925 | 941 | 814 |

**Table No. 5:** Odds Values, P-Values & C.I for table no 04.

| Major SARS-COV2<br>Symtoms |  | Samples With Positive<br>V/S ORF+N+E<br>Genes |  |  |  | Results |
| --- | --- | --- | --- | --- | --- | --- |
| 1 | COUGH | 0.83 | 0.015 | 0.72 | 0.96 | Weak<br>Association |
| 2 | SHORTNESS OF<br>BREATH/ DIFFICULTY<br>IN BREATHING | 0.7 | 0.00 | 0.61 | 0.81 | Weak<br>Association |
| 3 | Fever/ Chills | 2.66 | 0.00 | 2.3 | 3.07 | Strong<br>Association |
| 4 | NEW MUSCLE/<br>BODY ACHE | 3.4 | 0.00 | 2.91 | 3.97 | Strong<br>Association |
| 5 | SORE THROAT | 0.8 | 0.00 | 0.73 | 0.88 | Weak<br>Association |
| 6 | LOSS OF SMELL &<br>TASTE | 0.198 | 0.00 | 0.33 | 0.41 | Weak<br>Association |
| 7 | DIARRHEA | 0.18 | 0.00 | 0.15 | 0.21 | Weak<br>Association |
| 8 | HEAD ACHE | 0.024 | 0.00 | 0.02 | 0.034 | Weak<br>Association |
| 9 | NAUSEA | 1.29 | 0.00 | 1.12 | 1.48 | Strong<br>Association |
| 10 | VOMITTING | 1.85 | 0.00 | 1.61 | 2.13 | Strong<br>Association |
| 11 | RUNNY NOSE | 6.35 | 0.00 | 5.42 | 7.45 | Strong<br>Association |
| 12 | PERSISTANT CHEST<br>PAIN | 1.97 | 0.00 | 1.71 | 2.28 | Strong<br>Association |
| 13 | LAZINESS | 0.18 | 0.00 | 0.15 | 0.216 | Weak<br>Association |
| 14 | BLUISH LIPS/ FACE | 0.64 | 0.00 | 0.56 | 0.74 | Weak<br>Association |
|  |  | O.R | P-Value | LCI | UCI |  |

The presence of all three SARS-COV 2 genetic markers (i.e. ORF + N + E) was strongly associated with certain generalized/ nonspecific COVID-19 signs & symptoms like **Fever/Chills, New Muscular/Bodily Ache, Nausea, Vomiting & Persistent chest pain**.

### STRENGTH OF ASSOCIATION BETWEEN GENRALIZED/NON-SPECIFIC COVID-19 SINGS AND SYMPTOMS REPORTED BY ALL STUDY PARTICIPANTS WHOSE RT-PCR TEST WAS POSITIVE FOR DIFFERENT COMBINATIONS OF TWO SARS-COV 2 GENES (*i*.*e. ORF, N, E*) IDENTIFIED DURING RT-PCR PROCESS COMPARE TO OTHERS

As previously mentioned during the RT-PCR analysis process among all the 3375 study participants ***<u>only THREE</u>*** different (ORF, N, E) paired combinations were identified as summarized below:

**Table No. 6:** Subclasses of Study participants with paired ORF, N, E genes Combinations Identified during RT-PCR.

| ORF Gene | N Gene | E Gene | Total Number of Study Participants identified with this Combination | Percentage among all 3375 Study Participants |
| --- | --- | --- | --- | --- |
| - | + | + | 405 | 12% |
| + | + | - | 438 | 13% |
| + | - | + | 337 | 10% |

Over all 3375 study participants were sub divided into three different classes the nasopharyngeal and throat samples of these study participant only yields a pair of 2 genes out of all the three major genes that is ORF, N, & E gene. One class of study participants were identified with only N & E genes. Similarly, one class of study participants were positive only with ORF & N gene while some of the study participants only express ORF & E genes on to their RT-PCR report. The details of each sub class along with their numbers and percentages are shown in the above table.

**Table No. 7:** Contingency 2×2 table between generalized/ non-specific COVID-19 Signs and symptoms reported by all the study participants whose RT-PCR report was positive for different combinations of two SARS-COV 2 Genes (i.e. ORF, N, E) Compare to others.

| Different Combinations of 2 major Sars-Cov-2 Genes as Identified |  |  |  | Cough |  | O.R | C.I | P-value |
| --- | --- | --- | --- | --- | --- | --- | --- | --- |
| ORF | N | E |  | Yes | No |  |  |  |
| - | + | + | yes | 37 | 368 | 0.09 | 0.06 - 0.13 | 0.00 |
|  |  |  | No | 401 | 374 |  |  |  |
| + | + | - | Yes | 158 | 280 | 0.93 | 0.72 - 1.18 | 0.56 |
|  |  |  | No | 280 | 462 |  |  |  |
| + | - | + | Yes | 243 | 94 | 8.5 | 6.44 - 11.44 | 0.00 |
|  |  |  | No | 195 | 648 |  |  |  |

“Cough” was found to be strongly associated with the Genetic combination of (ORF + E) genes.
| Different Combinations of 2 major Sars-Cov-2 Genes as Identified |  |  |  | Fever/Chill |  | O.R | C.I | P-value |
| --- | --- | --- | --- | --- | --- | --- | --- | --- |
| ORF | N | E |  | Yes | No |  |  |  |
| - | + | + | yes | 258 | 147 | 1.7 | 1.3 - 2.28 | 0.00 |
|  |  |  | No | 384 | 391 |  |  |  |
| + | + | - | Yes | 223 | 215 | 0.79 | 0.63 - 1.01 | 0.06 |
|  |  |  | No | 419 | 323 |  |  |  |
| + | - | + | Yes | 161 | 176 | 0.68 | 0.53 - 0.88 | 0.003 |
|  |  |  | No | 481 | 362 |  |  |  |

“Fever/Chill” was found to be strongly associated with the Genetic combination of (N + E) genes.
| Different Combinations of 2 major Sars-Cov-2 Genes as Identified |  |  |  | Loss of smell & Taste |  | O.R | C.I | P-value |
| --- | --- | --- | --- | --- | --- | --- | --- | --- |
| ORF | N | E |  | Yes | No |  |  |  |
| - | + | + | yes | 351 | 54 | 2.62 | 1.89 - 3.63 | 0.00 |
|  |  |  | No | 552 | 223 |  |  |  |
| + | + | - | Yes | 269 | 169 | 0.27 | 0.20 - 0.35 | 0.00 |
|  |  |  | No | 634 | 108 |  |  |  |
| + | - | + | Yes | 283 | 54 | 1.88 | 1.35 - 2.61 | 0.00 |
|  |  |  | No | 620 | 223 |  |  |  |

“Loss of Smell & Taste” was found to be strongly associated with the Genetic combination of (N + E) genes.
| Different Combinations of 2 major Sars-Cov-2 Genes as Identified |  |  |  | Diarrhoea |  | O.R | C.I | P-value |
| --- | --- | --- | --- | --- | --- | --- | --- | --- |
| ORF | N | E |  | Yes | No |  |  |  |
| - | + | + | yes | 136 | 269 | 0.28 | 0.21 - 0.36 | 0.00 |
|  |  |  | No | 497 | 278 |  |  |  |
| + | + | - | Yes | 377 | 61 | 11.73 | 8.60 - 15.99 | 0.00 |
|  |  |  | No | 256 | 486 |  |  |  |
| + | - | + | Yes | 120 | 217 | 0.35 | 0.27 - 0.46 | 0.00 |
|  |  |  | No | 513 | 330 |  |  |  |

“Diarrhea” was found to be strongly associated with the Genetic combination of (ORF + N) genes.
| Different Combinations of 2 major Sars-Cov-2 Genes as Identified |  |  |  | Shortness of Breath / Difficulty in Breathing |  | O.R | C.I | P-value |
| --- | --- | --- | --- | --- | --- | --- | --- | --- |
| ORF | N | E |  | Yes | No |  |  |  |
| - | + | + | yes | 49 | 356 | 0.15 | 0.10 - 0.21 | 0.00 |
|  |  |  | No | 368 | 407 |  |  |  |
| + | + | - | Yes | 109 | 329 | 0.46 | 0.35 - 0.60 | 0.00 |
|  |  |  | No | 308 | 434 |  |  |  |
| + | - | + | Yes | 259 | 78 | 14.3 | 10.59 - 19.56 | 0.00 |
|  |  |  | No | 158 | 685 |  |  |  |

| Different Combinations of 2 major Sars-Cov-2 Genes as Identified |  |  |  | New Muscle/Body Ache |  | O.R | C.I | P-value |
| --- | --- | --- | --- | --- | --- | --- | --- | --- |
| ORF | N | E |  | Yes | No |  |  |  |
| - | + | + | yes | 175 | 230 | 0.54 | 0.42 - 0.68 | 0.00 |
|  |  |  | No | 453 | 322 |  |  |  |
| + | + | - | Yes | 146 | 292 | 0.26 | 0.21 - 0.34 | 0.00 |
|  |  |  | No | 482 | 260 |  |  |  |
| + | - | + | Yes | 307 | 30 | 16.64 | 11.15 - 24.82 | 0.00 |
|  |  |  | No | 321 | 522 |  |  |  |

“New Muscles/Body Ache” were found to be strongly associated with the Genetic combination of (ORF+ E) genes.
| Different Combinations of 2 major Sars-Cov-2 Genes as Identified |  |  |  | Sore Throat |  | O.R | C.I | P-value |
| --- | --- | --- | --- | --- | --- | --- | --- | --- |
| ORF | N | E |  | Yes | No |  |  |  |
| - | + | + | yes | 154 | 251 | 1.4 | 1.09 - 1.81 | 0.007 |
|  |  |  | No | 235 | 540 |  |  |  |
| + | + | - | Yes | 16 | 422 | 0.03 | 0.02 - 0.06 | 0.00 |
|  |  |  | No | 373 | 369 |  |  |  |
| + | - | + | Yes | 219 | 118 | 7.34 | 5.55 - 9.72 | 0.00 |
|  |  |  | No | 170 | 673 |  |  |  |

“Sore Throat” was found to be strongly associated with the Genetic combination of (ORF + E) genes.
| Different Combinations of 2 major Sars-Cov-2 Genes as Identified |  |  |  | Head Ache |  | O.R | C.I | P-value |
| --- | --- | --- | --- | --- | --- | --- | --- | --- |
| ORF | N | E |  | Yes | No |  |  |  |
| - | + | + | yes | 283 | 122 | 4.33 | 3.34 - 5.61 | 0.00 |
|  |  |  | No | 270 | 505 |  |  |  |
| + | + | - | Yes | 80 | 358 | 0.12 | 0.09 - 0.16 | 0.00 |
|  |  |  | No | 473 | 269 |  |  |  |
| + | - | + | Yes | 190 | 147 | 1.70 | 1.32 - 2.20 | 0.00 |
|  |  |  | No | 363 | 480 |  |  |  |

“Head ache” was found to be strongly associated with the Genetic combination of (N+E) genes.
| Different Combinations of 2 major Sars-Cov-2 Genes as Identified |  |  |  | Nausea |  | O.R | C.I | P-value |
| --- | --- | --- | --- | --- | --- | --- | --- | --- |
| ORF | N | E |  | Yes | No |  |  |  |
| - | + | + | yes | 97 | 308 | 0.12 | 0.09 - 0.15 | 0.00 |
|  |  |  | No | 560 | 215 |  |  |  |
| + | + | - | Yes | 323 | 115 | 3.43 | 2.65 - 4.45 | 0.00 |
|  |  |  | No | 334 | 408 |  |  |  |
| + | - | + | Yes | 237 | 100 | 2.38 | 1.82 - 3.12 | 0.00 |
|  |  |  | No | 420 | 423 |  |  |  |

“Nausea” was found to be strongly associated with Genetic combination of (ORF + N) genes.
| Different Combinations of 2 major Sars-Cov-2 Genes as Identified |  |  |  | Vomiting |  | O.R | C.I | P-value |
| --- | --- | --- | --- | --- | --- | --- | --- | --- |
| ORF | N | E |  | Yes | No |  |  |  |
| - | + | + | yes | 310 | 95 | 18.72 | 13.82 - 25.36 | 0.00 |
|  |  |  | No | 115 | 660 |  |  |  |
| + | + | - | Yes | 102 | 336 | 0.39 | 0.30 - 0.51 | 0.00 |
|  |  |  | No | 323 | 419 |  |  |  |
| + | - | + | Yes | 13 | 324 | 0.04 | 0.02 - 0.07 | 0.00 |
|  |  |  | No | 412 | 431 |  |  |  |

“Vomiting” was found to be strongly associated with Genetic combination of (N + E) genes.
| Different Combinations of 2 major Sars-Cov-2 Genes as Identified |  |  |  | Runny Nose |  | O.R | C.I | P-value |
| --- | --- | --- | --- | --- | --- | --- | --- | --- |
| ORF | N | E |  | Yes | No |  |  |  |
| - | + | + | yes | 91 | 314 | 0.27 | 0.21 - 0.36 | 0.00 |
|  |  |  | No | 397 | 378 |  |  |  |
| + | + | - | Yes | 199 | 239 | 1.30 | 1.02 - 1.65 | 0.02 |
|  |  |  | No | 289 | 453 |  |  |  |
| + | - | + | Yes | 198 | 139 | 2.71 | 2.09 - 3.52 | 0.00 |
|  |  |  | No | 290 | 553 |  |  |  |

“Runny Nose” was found to be strongly associated with Genetic combination of (ORF + E) genes.
| Different Combinations of 2 major Sars-Cov-2 Genes as Identified |  |  |  | Persistent Chest Pain Or Pressure |  | O.R | C.I | P-value |
| --- | --- | --- | --- | --- | --- | --- | --- | --- |
| ORF | N | E |  | Yes | No |  |  |  |
| - | + | + | yes | 375 | 30 | 21.73 | 14.5 - 32.40 | 0.00 |
|  |  |  | No | 283 | 492 |  |  |  |
| + | + | - | Yes | 118 | 320 | 0.13 | 0.10 - 0.18 | 0.00 |
|  |  |  | No | 540 | 202 |  |  |  |
| + | - | + | Yes | 165 | 172 | 0.68 | 0.52 - 0.87 | 0.00 |
|  |  |  | No | 493 | 350 |  |  |  |

“Persistent Chest pain/Pressure” was found to be strongly associated with Genetic combination of (N + E) genes.
| Different Combinations of 2 major Sars-Cov-2 Genes as Identified |  |  |  | Laziness |  | O.R | C.I | P-value |
| --- | --- | --- | --- | --- | --- | --- | --- | --- |
| ORF | N | E |  | Yes | No |  |  |  |
| - | + | + | yes | 145 | 260 | 0.58 | 0.45 - 0.74 | 0.00 |
|  |  |  | No | 379 | 396 |  |  |  |
| + | + | - | Yes | 340 | 98 | 10.52 | 7.95 - 13.91 | 0.00 |
|  |  |  | No | 184 | 558 |  |  |  |
| + | - | + | Yes | 39 | 298 | 0.09 | 0.06 - 0.13 | 0.00 |
|  |  |  | No | 485 | 358 |  |  |  |

“Laziness” was found to be strongly associated with Genetic combination of (ORF + N) genes.
| Different Combinations of 2 major Sars-Cov-2 Genes as Identified |  |  |  | Bluish Lips/Face |  | O.R | C.I | P-value |
| --- | --- | --- | --- | --- | --- | --- | --- | --- |
| ORF | N | E |  | Yes | No |  |  |  |
| - | + | + | yes | 209 | 196 | 0.42 | 0.32 - 0.54 | 0.00 |
|  |  |  | No | 555 | 220 |  |  |  |
| + | + | - | Yes | 294 | 144 | 1.18 | 0.92 - 1.51 | 0.18 |
|  |  |  | No | 470 | 272 |  |  |  |
| + | - | + | Yes | 261 | 76 | 2.32 | 1.73 - 3.10 | 0.00 |
|  |  |  | No | 503 | 340 |  |  |  |

“Bluish lips/Face” was found to be strongly associated with Genetic combination of (ORF + E) genes.

### STRENGTH OF ASSOCIATION BETWEEN GENRALIZED/NON-SPECIFIC COVID-19 SINGS AND SYMPTOMS REPORTED BY ALL STUDY PARTICIPANTS WHOSE RT-PCR TEST WAS POSITIVE FOR A SINGLE SARS-COV 2 GENES (*i*.*e. ORF or N or E*) COMPARE TO OTHERS

The following two tables summarizes the overall findings as shown below:

**Table No. 8:** Contingency 2×2 table between generalized/ non-specific COVID-19 Signs and symptoms reported by all the study participants whose RT-PCR report was positive for a single SARS-COV 2 Genes (i.e. ORF/ N/ E) Compare to others.

|  |  | COUGH |  | SHORTNESSS OF BREATH/DIFFICULTY IN BREATHING |  | Fever/Chills |  | NEW MUSCLE/ BODY ACHE |  | SORE THROAT |  | LOSS OF SMALL & TASTE |  | DIARRHEA |  |
| --- | --- | --- | --- | --- | --- | --- | --- | --- | --- | --- | --- | --- | --- | --- | --- |
|  |  | yes | No | yes | No | yes | No | yes | No | yes | No | yes | No | yes | No |
| ORF | + | 50 | 220 | 217 | 53 | 148 | 122 | 144 | 126 | 106 | 164 | 9 | 261 | 203 | 67 |
|  | - | 1088 | 2017 | 1187 | 1918 | 1968 | 1137 | 2138 | 967 | 1054 | 2051 | 1380 | 1725 | 1030 | 2075 |
|  |  | HEAD ACHE |  | NAUSEA |  | VOMITTING |  | RUNNY NOSE |  | PERSISTANT CHEST PAIN |  | LAZINESS |  | BLUISH LIPS/ FACE |  |
|  |  | yes | No | yes | No | yes | No | yes | No | yes | No | yes | No | yes | No |
| ORF | + | 160 | 110 | 35 | 235 | 209 | 61 | 102 | 168 | 136 | 134 | 96 | 174 | 21 | 249 |
|  | - | 778 | 2327 | 1607 | 1498 | 1611 | 1494 | 1986 | 1119 | 1959 | 1146 | 962 | 2143 | 1615 | 1490 |
|  |  | COUGH |  | SHORTNESSS OF BREATH/DIFFICULTY IN BREATHING |  | Fever/Chills |  | NEW MUSCLE/ BODY ACHE |  | SORE THROAT |  | LOSS OF SMALL & TASTE |  | DIARRHEA |  |
|  |  | yes | No | yes | No | yes | No | yes | No | yes | No | yes | No | yes | No |
| N | + | 57 | 146 | 71 | 132 | 50 | 153 | 160 | 43 | 52 | 151 | 109 | 94 | 18 | 185 |
|  | - | 1081 | 2091 | 1333 | 1839 | 2066 | 1106 | 2122 | 1050 | 1108 | 2064 | 1280 | 1892 | 1215 | 1957 |
|  |  | HEAD ACHE |  | NAUSEA |  | VOMITTING |  | RUNNY NOSE |  | PERSISTANT CHEST PAIN |  | LAZINESS |  | BLUISH LIPS/ FACE |  |
|  |  | yes | No | yes | No | yes | No | yes | No | yes | No | yes | No | yes | No |
| N | + | 114 | 89 | 73 | 130 | 135 | 68 | 132 | 71 | 160 | 43 | 154 | 49 | 120 | 83 |
|  | - | 824 | 2348 | 1569 | 1603 | 1685 | 1487 | 1956 | 1216 | 1935 | 1237 | 904 | 2268 | 1516 | 1656 |
|  |  | COUGH |  | SHORTNESSS OF BREATH/DIFFICULTY IN BREATHING |  | Fever/Chills |  | NEW MUSCLE/ BODY ACHE |  | SORE THROAT |  | LOSS OF SMALL & TASTE |  | DIARRHEA |  |
|  |  | yes | No | yes | No | yes | No | yes | No | yes | No | yes | No | yes | No |
| E | + | 80 | 22 | 95 | 7 | 73 | 29 | 40 | 62 | 94 | 8 | 11 | 91 | 95 | 7 |
|  | - | 1058 | 2215 | 1309 | 1962 | 2043 | 1230 | 2242 | 1031 | 1066 | 2207 | 1378 | 1895 | 1138 | 2135 |
|  |  | HEAD ACHE |  | NAUSEA |  | VOMITTING |  | RUNNY NOSE |  | PERSISTANT CHEST PAIN |  | LAZINESS |  | BLUISH LIPS/ FACE |  |
|  |  | yes | No | yes | No | yes | No | yes | No | yes | No | yes | No | yes | No |
| E | + | 70 | 32 | 35 | 67 | 49 | 53 | 28 | 74 | 24 | 100 | 56 | 46 | 36 | 66 |
|  | - | 868 | 2405 | 1607 | 1666 | 1771 | 1502 | 2060 | 1213 | 2071 | 1180 | 1002 | 2271 | 1600 | 1673 |

**Table No. 9:**
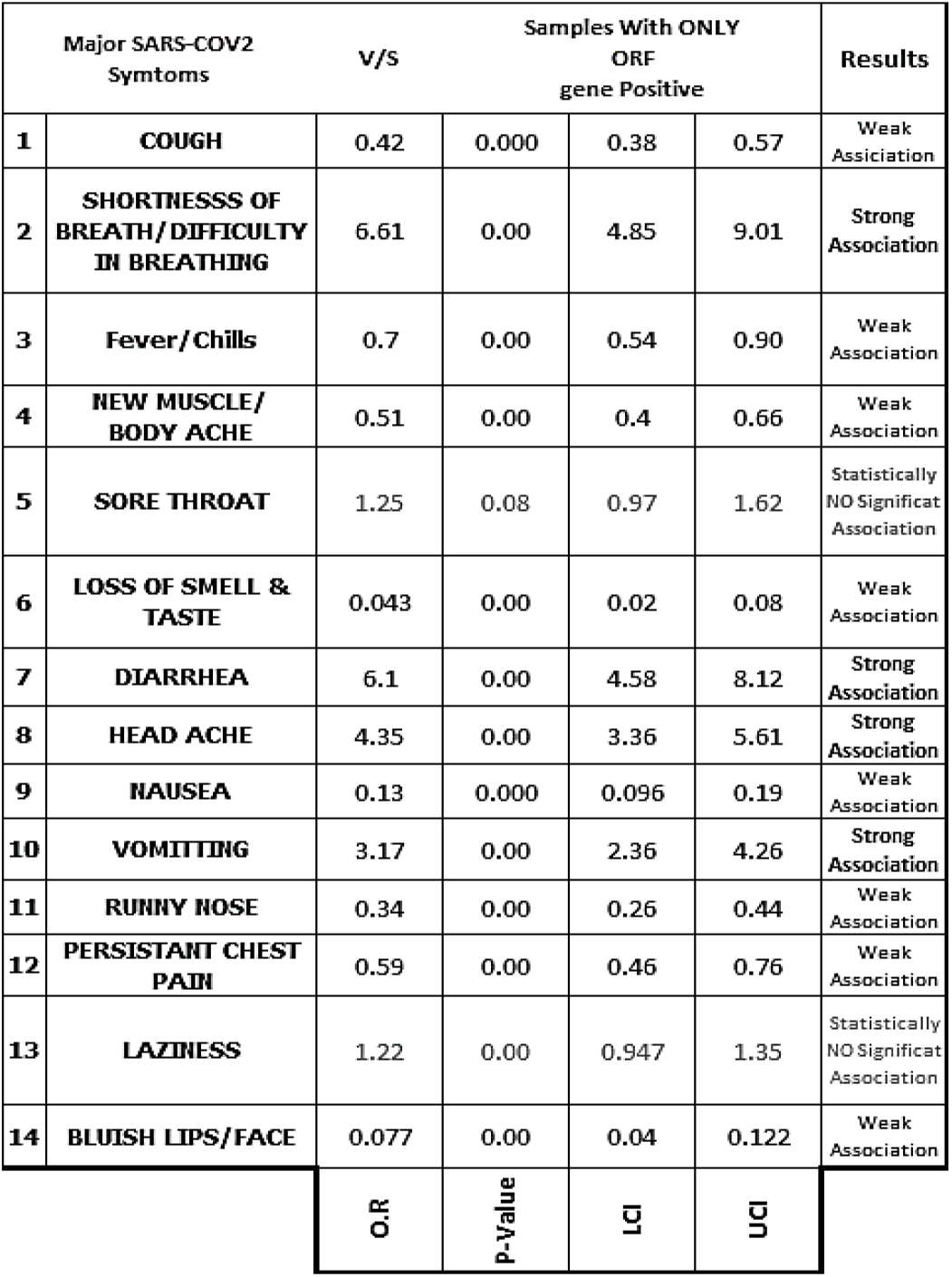
Odds Values, P-Values & C.I for table no 08.

**Table No. 10:**
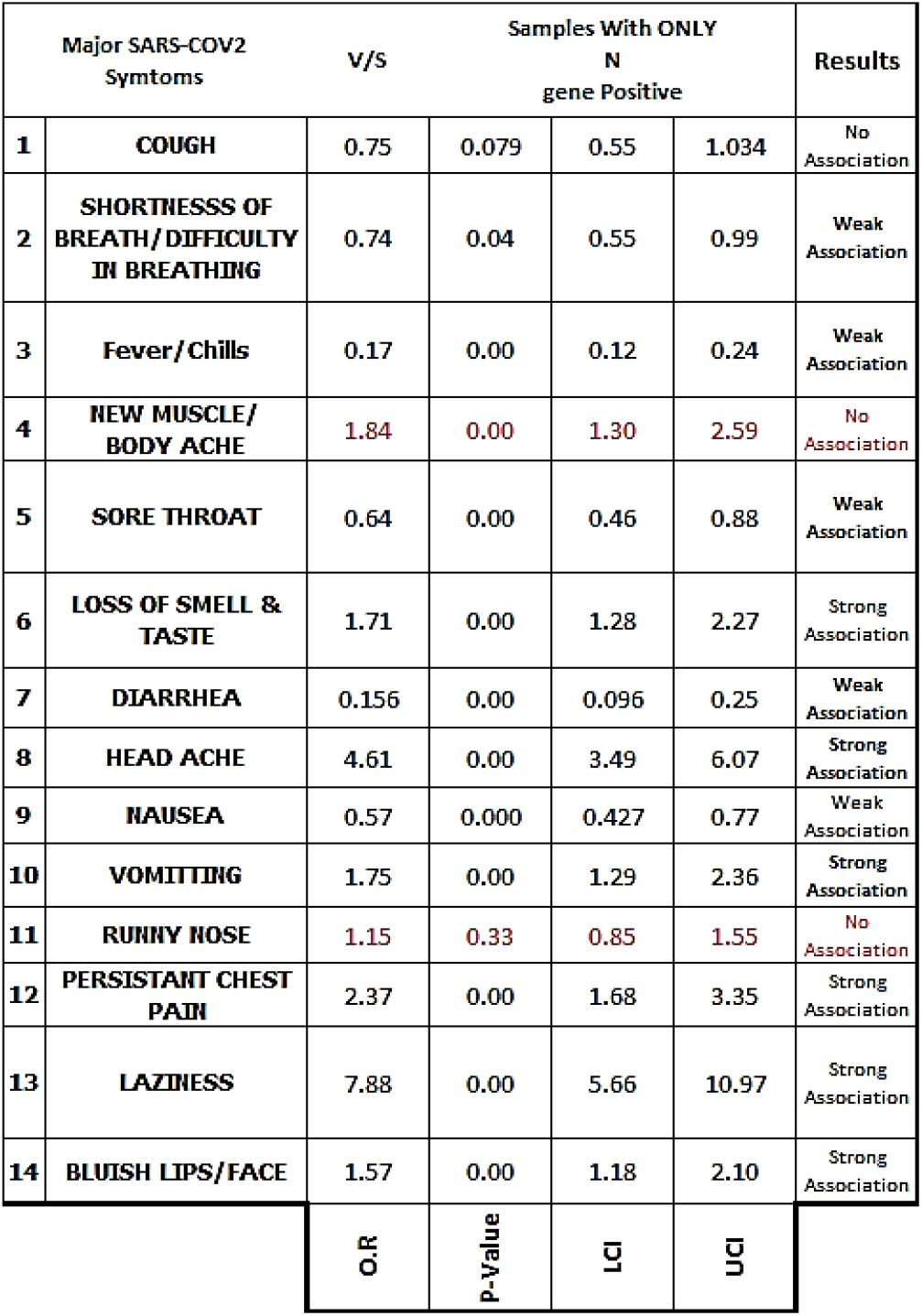
Odds Values, P-Values & C.I for table no 08.

The presence of **ORF gene** was found to be strongly associated “ Shortness of **Breath/Difficulty in Breathing”, Diarrhea, Head ache & Vomitting** while **Laiziness** was found to have no association with the presence of ORF gene. Similarly the presence of **N-gene** was found to be strongly associated with **Loss of smell & taste, Head ache**,**Presistant Chest Pain & Bluish lips/Face**, N-gene was found to have no association with **cough, New Muscular/Body ache & Runny Nose**.

**Table No. 11:** Odds Values, P-Values & C.I for table no 08.

| Major SARS-COV2 Symptoms |  | V/S | Samples With ONLY E gene Positive |  |  | Results |
| --- | --- | --- | --- | --- | --- | --- |
| 1 | COUGH | 7.61 | 0.000 | 4.72 | 12.27 | Strong Association |
| 2 | SHORTNESS OF BREATH/DIFFICULTY IN BREATHING | 20.76 | 0.000 | 9.61 | 44.86 | Strong Association |
| 3 | Fever/Chills | 1.51 | 0.00 | 0.98 | 2.34 | No Association |
| 4 | NEW MUSCLE/ BODY ACHE | 0.29 | 0.00 | 0.19 | 0.44 | Weak Association |
| 5 | SORE THROAT | 24.32 | 0.00 | 11.77 | 50.25 | Strong Association |
| 6 | LOSS OF SMELL & TASTE | 0.16 | 0.00 | 0.08 | 0.32 | Weak Association |
| 7 | DIARRHEA | 25.46 | 0.00 | 11.77 | 55.04 | Strong Association |
| 8 | HEAD ACHE | 6.06 | 0.00 | 3.96 | 9.27 | Strong Association |
| 9 | NAUSEA | 0.54 | 0.000 | 0.350 | 0.81 | Weak Association |
| 10 | VOMITTING | 0.78 | 0.22 | 0.52 | 1.16 | No Association |
| 11 | RUNNY NOSE | 0.22 | 0.00 | 0.143 | 0.34 | Weak Association |
| 12 | PERSISTANT CHEST PAIN | 0.13 | 0.00 | 0.08 | 0.21 | Weak Association |
| 13 | LAZINESS | 2.75 | 0.00 | 1.85 | 4.10 | Strong Association |
| 14 | BLUISH LIPS/FACE | 0.57 | 0.00 | 0.37 | 0.86 | Weak Association |
|  |  | O.R | P-Value | LCI | UCI |  |

The presence of **E-gene** was strongly associated with **Cough, Shortness of breath/ Difficulty in breathing, Sore throat, Diarrhea, Head ache & Laziness** while the **E-gene** was found to have no association with **Fever/Chill & Vomiting**.

## DISCUSSION & CONCLUSION

Previously it has been well established from at least 23 different nations, with 26 different clinical presentations that Six symptoms—fever (58.66%), cough (54.52%), dyspnea (30.82%), malaise (29.75%), weariness (28.16%), and sputum/secretion (25.33%)—had a general prevalence more than or equal to 25%. Other prevalent symptoms included headache (12.17%), chest discomfort (11.49%), diarrhea (9.59%), sneezing (14.71%), sore throat (14.41%), rhinitis (14.29%), goosebumps (13.49%), dermatological signs (20.45%), anorexia (20.26%), myalgia (16.9%), and rhinitis. The manifestations of dermatology were only documented in one study. Hemoptysis was the least common indication or symptom (1.65%). The three most common symptoms in trials involving more than 100 patients were dyspnea (30.64%), cough (54.21%), and fever (57.93%) ^[1-13]^. The study participants of our study also reported similar generalized/ nonspecific COVID-19 signs & symptoms.

From the very first case the SARS-COV 2 infection, a continuous mutation is reported in the virus as a result of which new variants popup frequently hence the overall virulence, treatment resistance, replication modalities & transmissions rates are all changing regularly hence in these circumstances it is of great importance to frequently monitor the symptomology of COVID-19. Currently no published literature has assessed the relationship of COVID-19 symptomology (i.e. *COUGH, SHORTNESSS OF BREATH/DIFFICULTY IN BREATHING, Fever/Chills, NEW MUSCLE/BODY ACHE, SORE THROAT, LOSS OF SMELL & TASTE, DIARRHEA, HEAD ACHE, NAUSEA, VOMITTING, RUNNY NOSE, PERSISTANT CHEST PAIN, LAZINESS, BLUISH LIPS/FACE*) with the *<u>ORF, N & E</u>* genes which are few of the diagnostic markers assessed for SARS COV-2 infection assessed during real-time PCR test.

The study findings showed that the presence of all three SARS-COV 2 genetic markers **(i.e. ORF + N + E)** was strongly associated with certain generalized/ nonspecific COVID-19 signs & symptoms like **Fever/Chills, New Muscular/Bodily Ache, Nausea, Vomiting & Persistent chest pain**.

Similarly, pair combination of different SARS-COV 2 virus genes identified showed that **(ORF + E)** gene pair presence was strongly associated **Cough, Shortness of breath/Difficulty in breathing, New Muscular/Body pain, Sore throat, Runny nose and Bluish lips/face**. Moreover, the **(N + E)** pair was found to be strongly associated with **Loss of smell & Taste, Head ache, Vomiting & Persistent chest pain/pressure**. Furthermore, the **(ORF + N)** pairing was found to be strongly associated with **Diarrhea, Nausea & Laziness**.

Our study also showed that individually the presence of **ORF gene** was found to be strongly associated **“ Shortness of Breath/Difficulty in Breathing”, Diarrhea, Head ache & Vomitting** while **Laiziness** was found to have no association with the presence of ORF gene. Similarly the presence of **N-gene** was found to be strongly associated with **Loss of smell & taste, Head ache**,**Presistant Chest Pain & Bluish lips/Face**, N-gene was found to have no association with **cough, New Muscular/Body ache & Runny Nose**. While the presence of **E-gene** was strongly associated with **Cough, Shortness of breath/ Difficulty in breathing, Sore throat, Diarrhea, Head ache & Laziness** while the **E-gene** was found to have no association with **Fever/Chill & Vomiting**.

## Data Availability

Data can be shared upon request.

