## Supplementary 1 for "STRENGTH OF ASSOCIATION BETWEEN GENERALIZED/NONSPECIFIC COVID-19 SIGNS & SYMPTOMS WITH SARS-COV 2 SPECIFIC ORF, N, E GENES IDENTIFIED THROUGH REAL TIME PCR"

PROVINCIAL HEALTH DIRECTORATE, BALOCHISTAN, QUETTA.
**

**I.D # 122234/FELTP/2021** **Dated Quetta 1st January, 2021**

**TO WHOM IT MAY CONCERN--- (INSTITUTIONAL REVIEW BOARD) ---- DECISION.**

This supreme institutional review board under the chairmanship of ***"Director General Health Services Balochistan" have*** gone thoroughly through the research proposal submitted to us under the title of **"STRENGTH OF ASSOCIATION BETWEEN GENERALIZED/NONSPECIFIC COVID-19 SIGNS & SYMPTOMS WITH SARS-COV 2 SPECIFIC ORF, N, E GENES IDENTIFIED THROUGH REAL TIME PCR.**

- We found that this study proposal is totally an observational study, no humans or animals will be subjected to any sort of harm. Moreover for ethical consideration the guidelines of ***"World Medical Association's Declaration of Helsinki"*** were strictly considered during the review process and we found no contradiction to them.

- Hence the Institutional Review Board approves this study.

**Member #01:** *Dr. Sagheer Ahmed* **(Remarks:** Approved**).**
**Member # 02:** *Dr. Aslam Kakar* **(Remarks:** Approved**).**

**Dr. Nasir Ali Bugti.**
**Director General Health Services**
**Balochistan Quetta. (Chairman)**

**Copy to the:-**

1. Executive Director, National Institute of Health (NIH) Islamabad.
2. The Director Programs, MoNHSR&C Islamabad
3. The National FP IDSR NIH Islamabad
4. The MSDHQ Hospital Sibi &Zhob
5. The District Health Officer Sibi &Zhob
6. The Provincial IDSR FP/ Technical Support Officer PDSRU_FELTP Balochistan Quetta
7. The Chief Planning Officer Health Department Balochistan Quetta
8. PS to the Secretary Primary & Secondary Health Care Balochistan Quetta.

**Dr. Nasir Ali Bugti.**
**Director General Health Services**
**Balochistan Quetta.**
